# When do persuasive messages on vaccine safety steer COVID-19 vaccine acceptance and recommendations? Behavioral insights from a randomised controlled experiment in Malaysia

**DOI:** 10.1101/2022.04.17.22273942

**Authors:** Nicholas Yee Liang Hing, Yuan Liang Woon, Yew Kong Lee, Joon Kim Hyung, Nurhyikmah M. Lothfi, Elizabeth Wong, Komathi Perialathan, Nor Haryati Ahmad Sanusi, Affendi Isa, Chin Tho Leong, Joan Costa-Font

## Abstract

**Introduction:** Vaccine safety is a primary concern among vaccine hesitant individuals. We examined how seven persuasive messages with different frames, all focusing on vaccine safety, influenced Malaysians to accept the COVID-19 vaccine, and recommend it to individuals with different health and age profiles; i.e. healthy adults, elderly, and people with pre-existing health conditions.

**Methods:** A randomized controlled experiment was conducted among 5,784 Malaysians who were randomly allocated into 14 experimental arms. They were exposed to one or two messages that promoted COVID-19 vaccination. Interventional messages were applied alone or in combination and compared against a control message. Outcome measures were assessed as intent to both take the vaccine and recommend it to healthy adults, elderly, and people with pre-existing health conditions, before and after message exposure. Changes in intent after message exposure were modeled and we estimate the average marginal effects with respect to changes in the predicted probability of selecting a positive intent for all four outcomes.

**Results:** The average baseline proportion of participants with positive intents in each arm to take, and recommend the vaccine to healthy adults, elderly, and people with pre-existing health conditions was 61.6%, 84.9%, 72.7% and 51.4% respectively. We found that persuasive communication via several of the experimented messages improved recommendation intent to people with pre-existing health conditions, with improvements ranging between 4 to 8 percentage points. In contrast, none of the messages neither significantly improved vaccination intentions, nor recommendations to healthy adults and the elderly. Instead, we found evidence suggestive of backfiring among this group with messages using negative frames, risky choice frames, and priming descriptive norms.

**Conclusion:** Persuasive messages aimed at influencing vaccination decisions should incorporate a combination of factors linked to hesitancy. Messages intended to promote recommendation of novel health interventions to people with pre-existing health conditions should incorporate safety dimensions.

**Clinical Trials registration number:** NCT05244356

**Key Messages:** *What is already known?:*

- Persuasive messages have been shown to influence COVID-19 vaccination intentions, but evidence from Low- and Middle-income countries are limited.
- There are limited studies investigating the effect of persuasive messages in influencing decisions to recommend the COVID-19 vaccine, with only a single study to date which investigated effects directed at recommending the COVID-19 vaccine to a friend, but without considering the individual’s health and age profile.

*What are the new findings?:*

- Persuasive messages that focused on vaccine safety did not positively influence Malaysian adults to take the COVID-19 vaccine or recommend it to healthy adults and the elderly, while messages framed as descriptive norms, negative attribute, and risky choices, significantly backfired for some of these outcomes.
- Several persuasive messages focusing on vaccine safety significantly improved intent to recommend the COVID-19 vaccine to people with pre-existing health conditions.

*What do the new findings imply?:*

- Instead of only addressing safety, persuasive messages aimed at nudging vaccination should incorporate multiple behavioral determinants linked to vaccine acceptance.
- Persuasive messages that are intended to promote uptake of novel health interventions should incorporate safety dimensions as a form of assurance for others to recommend it to people with pre-existing health conditions, given that they may be perceived as more susceptible to hazards from adverse events.

## Introduction

The COVID-19 pandemic has sparked global efforts to develop counter measures against SARS-CoV-2. One such measure lies with the rapid research and development of effective COVID-19 vaccines[1] which are critical to achieve impactful COVID-19 vaccination campaigns.[2] Although credible vaccine information from official sources are abundantly available,[3–6] vaccine hesitant individuals risk compromising widespread vaccination[7] as they delay or refuse to take a vaccine once it is made available.[8]

Vaccine safety remains one of the top reasons cited by vaccine hesitant individuals in Malaysia and abroad.[9–12] This is aggravated by misinformation regarding COVID-19 vaccine safety.[13] Hence, a question that emerges is how best to effectively communicate vaccine safety information. A potential method stems from applying nudges, which alters the environmental or information context to encourage a particular behavior.[14] One such form involves using various frames of persuasive messages that employ nudge techniques to encourage behaviour change[15,16].

Few studies have experimented messages that specifically address COVID-19 vaccine safety among unvaccinated individuals. Persuasive messages emphasizing vaccine safety either through explaining the rigorous process of drug development, leveraging the authority of a clinician to explain vaccine safety, or highlighting vaccine approval from a regulatory agency, failed to significantly improve vaccination intent.[17–19] Conversely, positive effects were observed with risky choice framed messages that favorably juxtaposed vaccine risk versus contracting COVID-19.[20] However, the message was tested among employees of a healthcare organisation who may have higher health awareness compared to the general public. Barnes & Colagiuri also observed positive effects with messages applying attribute framing whereby vaccine side effect rates were framed positively or negatively.[21] However, they investigated booster shot intentions among fully vaccinated individuals.

Based on current available evidence, there are several knowledge gaps. The effects of attribute framing have yet to be explored among unvaccinated individuals. Risky choice frames that pit vaccine related death rates or side effects against the COVID-19 disease has not been studied among the general public. Although descriptive norms have been widely studied to nudge COVID-19 vaccination by invoking psychological judgments that vaccination is a societally approved behavior, [19,20,22,23] these messages have yet to be framed to imply vaccine safety as a motivation for vaccination among the majority. The effect of using vaccinated health authorities to imply vaccine safety and recommend the vaccine has also not been thoroughly studied. Finally, given that individual decision and behavior are intrinsically linked to context and culture,[24–26] there are reasons to believe that vaccination nudges ought to be adapted to Low- and Middle-income countries (LMIC) such as Malaysia. However, there exists a paucity of information for using such nudges in LMIC, with most published evidence originating from developed countries. [16]

Furthermore, previous studies have widely investigated nudges to influence personal interests to vaccinate one’s self or own child [16,27,28] rather than a person’s decision to recommend vaccination. Although James *et al* did investigate the effects of persuasive messages in recommending a COVID-19 vaccine to a friend, they did not consider the health or age profile of the person being recommended.[29] Having a finer gauge on which group of people have higher likelihoods to be recommended is important especially in Asian communities who pay special attention to advice sought from family and friends with significance when making a health related decision.[30]

Therefore, an experiment using various message frames intended to narrow the current knowledge gaps was conducted in Malaysia. Our primary objectives were to investigate whether persuasive messages focusing on vaccine safety influenced the intention to take the COVID-19 vaccine, and recommend it to healthy adults, the elderly who are aged 60 and above, and people with pre-existing health conditions. Apart from examining single messages, we investigate the effects of combining messages together to mimic a real-world environment where people are exposed to multiple messages. This allows us to determine if combining messages resulted in improved effectiveness from a higher message dose effect,[31] or reduced effectiveness due to message interactions causing a boomerang effect.

## Methods

### Study design

We conducted a prospective 14-arm randomised controlled experiment with a parallel design. The experiment was conducted using a web-based survey that was launched on a platform belonging to Dynata, an international market research company based in America. The company has an online survey panel composed of Malaysians who have signed up on the survey platform. Participants who complete a survey will receive reward points as per Dynata’s policy.

### Study participants and setting

The experimental survey was conducted from the 29^th^ of April to 7^th^ of June 2021 (more details about the COVID-19 situation in Malaysia during participant recruitment can be found in the online supplemental material). Participants were recruited from Dynata’s online survey panel. Eligible participants were adult Malaysians aged 18 years and above who could understand either the English or Malay language and had not received any dose of the COVID-19 vaccine. The latter criterion was included as the majority of the population has not been vaccinated during the experimental period. Additionally, this criterion allowed us to investigate the effectiveness of messages in an unvaccinated population, which is an important aspect prior to any novel vaccination roll-out.

Sample size requirement was calculated based on a logistic model to detect a small effect size of 0.1, with the baseline proportion of people who definitely will take the COVID-19 vaccine set at 0.67. This baseline value was chosen based on the reported proportion of Malaysians willing to accept the COVID-19 vaccine in a national survey that was conducted before this study was being planned.[12] Sample size was calculated to be 318 respondents per arm, after setting power at 80% and significance level at 0.05. Taking into account a 20% drop out rate in the event of invalid responses, calculated sample size was 400 participants per arm. Participants were recruited via stratified sampling based on age, gender, ethnicity and household income to obtain an approximately population-representative sample (more details about the stratified sampling can be found in the online supplemental material).

All participants were shown a page that described background information about the study before they provided informed consent by clicking on a button indicating agreement to join the experiment. This experiment received ethical approval from the Medical Research Ethics Committee of the Malaysian Ministry of Health.

### Randomisation and masking

Enrolled participants were randomly allocated into a particular experimental arm by Dynata through an automated computer randomization system. This experiment was double blinded whereby participants were unaware of what interventional message was given to them and investigators had no control over treatment assignment as this aspect was completely handled by the market research company.

### Data collection and intervention

Sociodemographic variables that screened for inclusion criteria and enabled stratified sampling during experimental arm allocation were first collected from approached participants. Recruited participants were measured on their general attitude towards vaccines as this factor has been shown to significantly influence vaccine uptake intent.[28,32] Attitude was assessed by measuring the level of agreement; using a five-point Likert scale, of two statements regarding the efficacy of vaccines in protecting against serious diseases, and personal religious or cultural backing for vaccination. Participants were also asked in the remaining sociodemography section, whether they had refused towards vaccinating their child in the past. These questions were adapted from locally conducted studies.[32,33] Participants were categorised as having potential negative attitudes if they provided responses indicating disagreement, uncertainty or refusal to any of those questions.

Participants were then asked a series of questions relating to their baseline intentions to accept and recommend the COVID-19 vaccine before being randomly assigned to an experimental arm. Participants were exposed to either one or two messages from a selection of 8 different types of messages and were instructed to read the message completely before clicking a button to proceed. Each message was calibrated to be on screen for at least eight seconds before participants could proceed to prevent them from skipping the message. Table 1 describes the content of each message and the corresponding nudge technique that the content was incorporated with. The source of the information displayed is stated below the message’s content to provide information credibility.

**Table 1:** Content of each experimental treatment message used along with the corresponding nudge technique employed. Each message is assigned a code to ease referencing.

| No. | Nudge technique | Content | Message code |
| --- | --- | --- | --- |
| 1 | Descriptive norm. | Around 70% of Malaysians said they will get the COVID-19 vaccine.<br>*Source:*<br>*Ministry of Health, Malaysia; 31 December 2020 [12]* | **DN(70%)** |
| 2 | Descriptive norm. | The COVID-19 vaccine was tested with thousands of people, including the elderly, and people with existing health conditions.<br>Now millions of people worldwide have received it.<br>When it's your turn, you can be confident that it is safe and effective.<br>*Source:*<br>*Kyriakidis et al. NPJ Vaccines. 2020 February 22;6(1):28 [34]*<br>*John Hopkins Coronavirus Resource Center, United States of America [35]* | **DN** |
| 3 | Influence from a Government official and health authority, and descriptive norm. | Malaysia's Health Director General, Dr. Noor Hisham Abdullah, and 9 out of 10 healthcare workers in Malaysia have received the COVID-19 vaccine.<br>They recommend that you get it too.<br>*Source:*<br>*COVID-19 Immunisation Task Force, Malaysia; 23 February, 29 March 2021 [36,37]* | **HCW** |
| 4 | Negative attribute framing. | Only 4 out of 100 people who received the COVID-19 vaccine experienced side effects.<br>*Source:*<br>*Ministry of Health, Malaysia; 2 April 2021 [38]* | **NF** |
| 5 | Positive attribute framing. | 996 out of 100 people who received the COVID-19 vaccine did not experience any side effects.<br>*Source:*<br>*Ministry of Health, Malaysia; 2 April 2021 [38]* | **PF** |
| 6 | Risky choice framing (Safety). | There are 0 deaths caused by the COVID-19 vaccines.<br>On the other hand, over 1,400 people have died due to COVID-19.<br>*Source:*<br>*Ministry of Health, Malaysia; 18 March 2021 [39]*<br>*Crisis Preparedness and Response Centre (CPRC), Malaysia; 23 April 2021 [40]* | **RC(S)** |
| 7 | Risky choice framing (Side effects). | Only 4 in 1 million people who received the COVID-19 vaccine experienced blood clots.<br>On the other hand, 200,000 in 1 million people infected with COVID-19 experienced blood clots.<br>*Source:*<br>*Torjesen I. The British Medical Journal. 2021;373:n1005 [41]*<br>*Malas et al. EClinicalMedicine. 2020 December; 29:100639. [42]* | **RC(SE)** |
| 8 | Control message. | Get the COVID-19 Vaccine.<br>It's safe and effective! | N/A |

The control message was devoid of any nudge element and only displays a slogan that rallies the reader to get the COVID-19 vaccine because it is safe and effective. The other experimental messages begin with an opening tagline highlighting the main concern that Malaysians have about the COVID-19 vaccine and serves as a precursor for the following message content which attempts to alleviate that concern. Each message concludes with a rally slogan that is identical with the control message. Each message was validated with at least five people and went through a series of iterations to ensure that the content was interpreted correctly (details about the message design and examples of actual messages can be found in the online supplemental material and supplemental figure S1 respectively).

Our experiment presents a total of 14 arms. Participants were exposed to one message in the first eight arms, and two messages in the remaining arms. *DN(70%)* exposure was held constant in arms bearing two messages. This message was made a constant because it focuses on Malaysians as the persona of interest, which makes it the most personally relevant message to our Malaysian participants. Participants who received two messages were exposed to each message one at a time, with the sequence of appearance being random.

After message exposure, participants were asked again regarding intentions to receive and recommend the COVID-19 vaccine. Participants who were hesitant about taking or recommending the vaccine post exposure were asked about the possible reasons for such responses. Lastly, remaining sociodemographic variables such as education level were collected.

### Outcomes

Intent to accept the COVID-19 vaccine was assessed by asking participants whether they would take the COVID-19 vaccine. Participants answered using a four-Likert scale which indicated “Definitely no”, “Not sure, but probably no”, “Not sure, but probably yes” and “Definitely yes”. This scale was used to eliminate subjective ambiguity and allows participants to express their intent in detail which is capably determined as it involves an internalised decision.[43]

Intent to recommend the COVID-19 vaccine was assessed by asking participants to rate their level of agreement on recommending the vaccine to three groups of family members namely, healthy adults, elderly, and members with any pre-existing health conditions. Family members were chosen as a target character because they are related to respondents, thus yielding unbiased responses regarding intent to recommend to each of the three studied groups. Participants rated their agreement via a five-Likert scale which provided options of “Definitely no”, “Probably no”, “Not sure”, “Probably yes”, and “Definitely yes”. This scale was chosen to provide a neutral answer in the form of a “Not sure” option, because the decision to recommend may influence the outcome of another individual, which may be a difficult decision and thus warrant a neutral stance.

Our four study objectives were based on outcomes measured at baseline and post intervention. Positive intent was defined as responding “Definitely yes” and “Agree” or “Strongly agree” for accepting and recommending the vaccine respectively. These responses indicated absolutely no hesitancy towards the action in question whereas the remaining options reflected uncertainty or refusal.

### Statistical analyses

Summary statistics (frequency and percentages, mean and standard deviation) of recruited participants’ demographics, attitude towards vaccines, and intent to accept and recommend the COVID-19 vaccine in each experimental arm, was reported. Balance tests were conducted to check if baseline characteristics were significantly different between each experimental arm.

Since the responses for all four outcome measures were ordinal in nature, we applied four separate generalised ordered logistic regressions to estimate how each experimental arm affected the propensity of selecting a particular level of intent. Each regression model was adjusted for general attitude towards vaccines and baseline intent that corresponds to the outcome measure analysed. Generated regression models were subsequently used to compute the average marginal effects of each interventional arm relative to the control arm with respect to changes in the predicted probability of selecting a positive intent for all four outcome measures. This provided an estimate behind the effectiveness and probability change magnitudes exerted by experimented messages.

Additionally, we tested heterogeneous treatment effects of age, gender, and education level to investigate whether our intervention messages impacted certain groups of individuals differently. All analyses were conducted using Stata version 16. This study was registered on clinicaltrials.gov (ID number: NCT05244356).

## Results

A total of 5,784 participants were recruited into the experiment. Each arm was assigned between 410 to 416 participants. Figure 1 presents the experimental design flow chart. Due to the experiment being an online survey, our sampled dataset was skewed towards younger participants and those who had pre-university or tertiary education. However, all experimental arms were balanced and showed no significant differences with respect to key baseline characteristics. The average baseline proportion of participants with positive intents in each arm to take, and recommend the COVID-19 vaccine to healthy adults, elderly, and people with health conditions, was 61.6%, 84.9%, 72.7% and 51.4% respectively. Summary statistics of survey participants stratified according to experimental arms can be found in online supplemental table S1.

**Figure 1:**
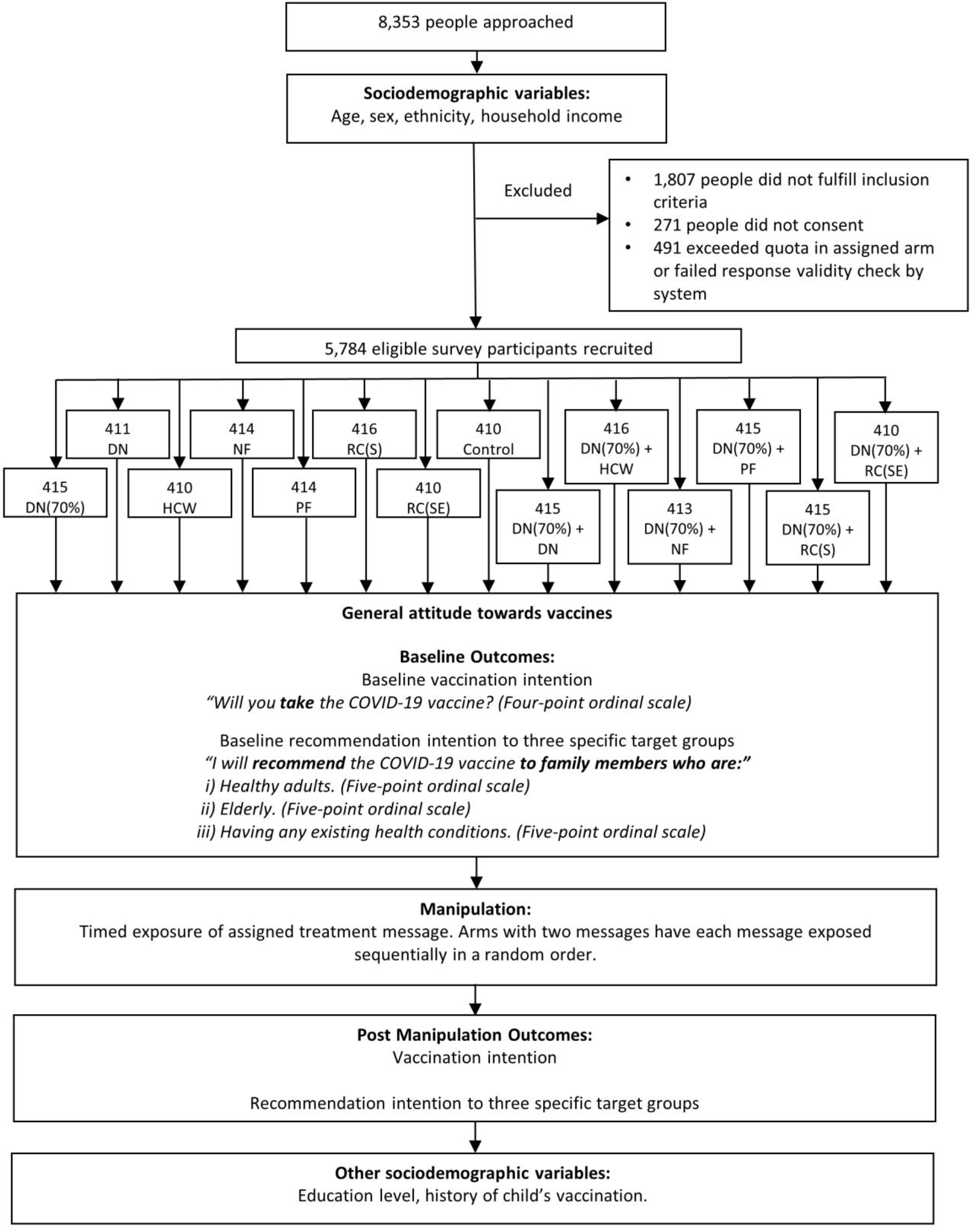
Experimental design flow chart presenting sample size, arm allocations, and item wordings for outcomes.

Figure 2 depicts forest plots that describe the average marginal effects of providing positive responses for each interventional arm relative to the control arm in all outcomes measured. A summary of marginal effects for all levels of responses can be found under online supplemental tables S2 and S3. In terms of participant’s intent to accept the COVID-19 vaccine, none of the interventional arms were significantly effective at improving intent compared to the control message. However, we found that intent to vaccinate significantly dropped by 3.3 percentage points (95% CI -6.3 to -0.2) among participants assigned to the *NF* message. Combining *DN(70%)* to *NF* had a transient added reduction in intent to vaccinate (3.5 percentage points, 95% CI -6.6 to - 0.5). None of the interventional arms were significantly effective at improving recommendation intent towards healthy adults and the elderly. Instead, recommendation intent towards healthy adults significantly fell in the *DN(70%)* and *RC(S)* arm by 3.8 percentage points (95% CI -7.0 to -0.7) and 4.3 percentage points (95% CI -7.5 to -1.0) respectively. Intent to recommend healthy adults in the combination message arm containing *DN* dropped by 3.3 percentage points (95% CI -6.4 to -0.1) whilst the arm with *RC(S)* reduced by 3.2 percentage points (95% CI -6.4 to -0.0). Conversely, five interventional arms were significantly effective at improving recommendation intent to people with pre-existing health conditions. The *DN* arm showed highest significant improvements at 8 percentage points (95% CI 4.1 to 12.0). The *PF* was the second-best performing arm, improving intent by 5.6 percentage points (95% CI 1.7 to 9.5). The remaining arms showing significant improvements were the combination messages containing *DN* (4.2 percentage points, 95% CI 0.2 to 8.1), *HCW* (4.7 percentage points, 95% CI 0.8 to 8.6), and *RC(S)* (4.6 percentage points, 95% CI 0.7 to 8.5) message.

**Figure 2:**
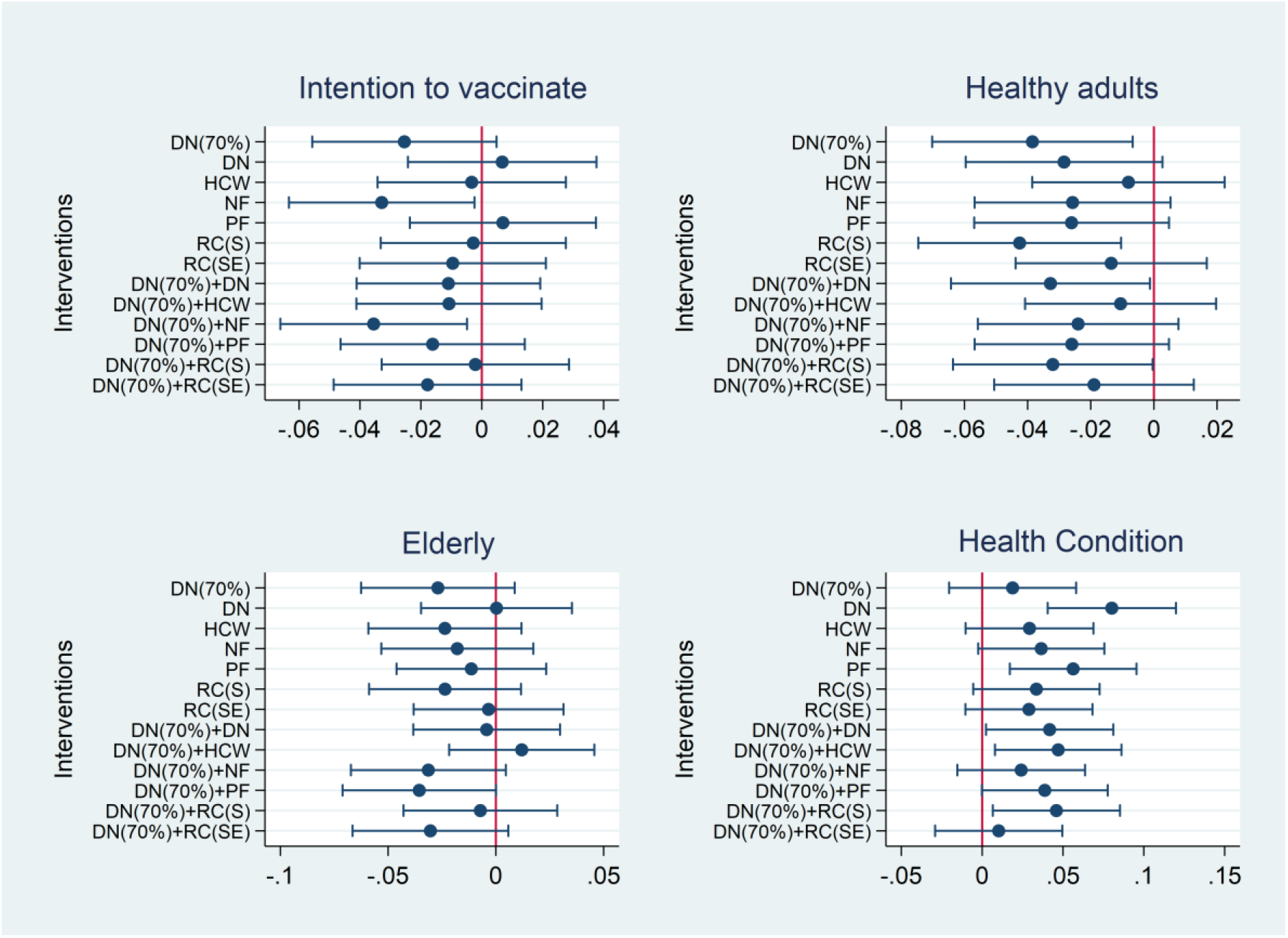
Average marginal effects of predicted probabilities for each interventional arm relative to the control arm for all primary outcome measures; i) Intention to vaccinate, ii) Recommend healthy adults (Healthy adults), iii) Recommend elderly (Elderly), iv) Recommend people with pre-existing health conditions (Health condition). Scatter plots present point estimates, 95% confidence intervals and the line of indifference.

Being worried about the safety or side effects of the vaccine was the main reason for hesitancy, with 70% to 80% participants who were hesitant in each outcome answering as such. A tabulation that shows the proportion of respondents citing reasons for hesitancy for each outcome can be found under online supplemental figures S2 to S5. We found no significant differences between all arms with respect to proportion of respondents citing this top reason (online supplemental table S4).

Figure 3 shows forest plots with 95% confidence intervals for heterogeneous treatment effects that indicate definite intentions of accepting the COVID-19 vaccine and agreeing to recommend it. A summary of treatment effect values can be found under online supplemental tables S5 to S7. There is evidence showing certain sociodemographic groups are more impacted by our experimented messages.

**Figure 3:**
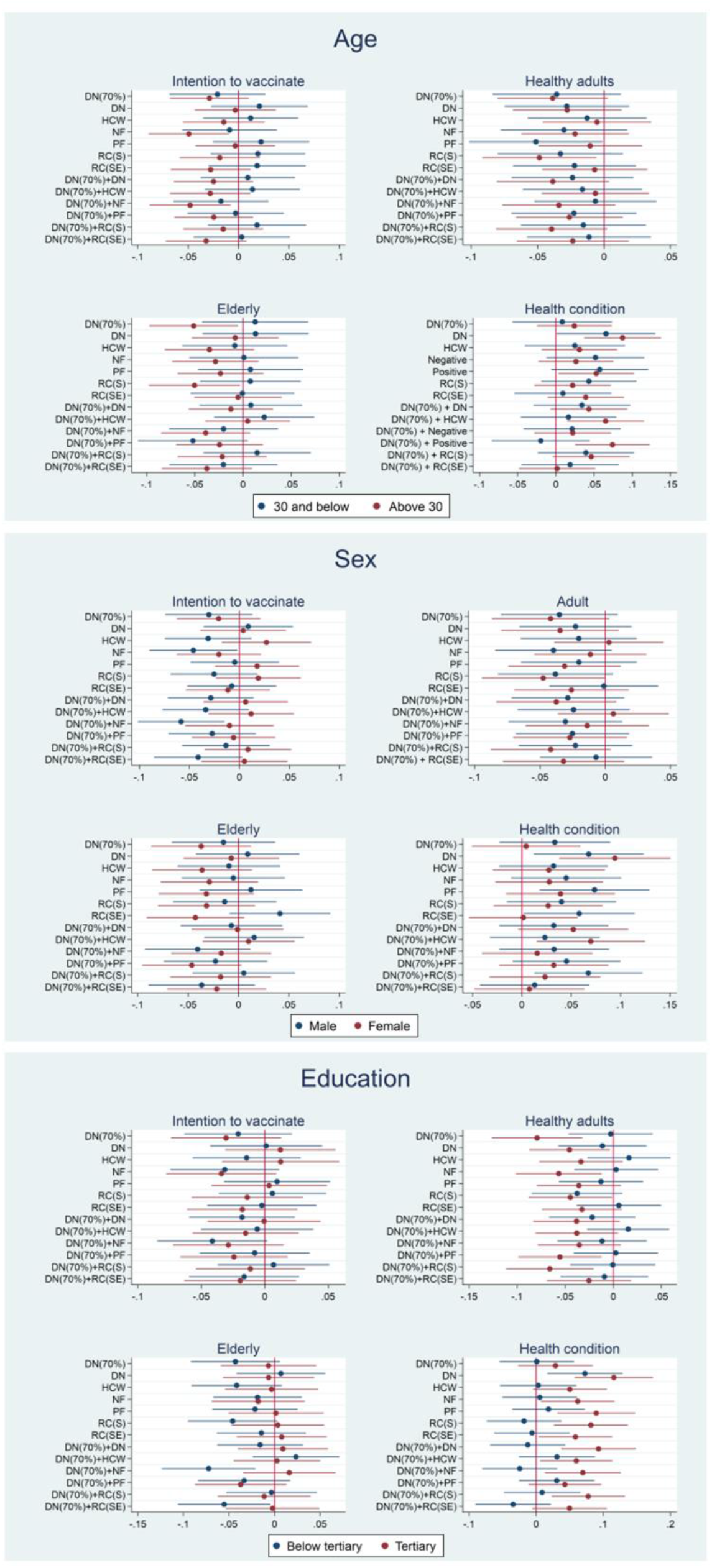
Sociodemographic determinants of average marginal treatment effects to select option of definitely intending to accept the vaccine and agreeing to recommend the vaccine based on age, sex and education for each treatment arm relative to control arm, with respect to all 4 primary outcome measures; i) Intention to vaccinate, ii) Recommend healthy adults (Healthy adults), iii) Recommend elderly (Elderly), iv) Recommend people with pre-existing health conditions (Health condition). Scatter plots present point estimates, 95% confidence intervals and the line of indifference.

Subgroup analysis for participants aged below and above 30 years old was conducted. This grouping was selected to investigate if youths, who have much lesser risk for suffering severe disease but have their future wellbeing affected,[44] responded differently compared to the older age groups who have higher risk for serious complications.[45] Our analyses show that only the older participants were significantly affected by the *NF* message when applied alone or when in combination with *DN(70%)*, registering a drop in intent by almost 5 percentage points. A similar drop in intent to recommend healthy adults was observed among the older age group and youths who were exposed to the *RC(S)* and *PF* message respectively. There was also an almost equal drop in intent to recommend the vaccine to elderly among older participants exposed to *DN(70%)* and *RC(S)*. Both age groups responded positively to the *DN* message for intent to recommend people with health conditions, whereby youths and older people saw an increase in intent by 6.6 percentage points (95% CI 0.1 to 13.0) and 8.7 percentage points (95% CI 3.7 to 13.8) respectively. Older people also showed an increase in intent to recommend by 5.3 percentage points (95% CI 0.3 to 10.3) when exposed to *PF*. Intent increased to 7.4 percentage points (95% CI 2.5 to 12.3) when *DN(70%)* was added. Additionally, older people were positively influenced to recommend when *DN(70%)* was combined with *HCW* (6.5 percentage points, 95% CI 1.5 to 11.5).

Between sexes, males were more negatively impacted by the *NF* message, having vaccination intent drop by 4.6 percentage points (95% CI -9.0 to -0.2). Vaccination intent further declined to 5.8 percentage points (95% CI -10.2 to -1.5) when *DN(70%)* was added. Females were more negatively influenced by the *RC(S)* message, causing a reduction in intent to recommend healthy adults by 4.8 percentage points (95% CI -9.5 to -0.0). However, both sexes had significantly increased intent to recommend people with health conditions when they were exposed to the *DN* message whereby intent improved by 6.8 percentage points (95% CI 1.2 to 12.3) and 9.4 percentage points (95% CI 3.8 to 15.1) for males and females respectively. Males were also more positively influenced by both *PF* and *RC(SE)* messages. Moreover, males tended to positively respond when *DN(70%)* was combined with *RC(S)* whilst females exhibited a similar response when *DN(70%)* was combined with *HCW*.

Subgroup analysis was conducted between participants with and without tertiary education to observe any differences in behavioral response to the messages, given that Malaysians with a bachelor’s degree or higher were more accepting of the COVID-19 vaccine.[46] None of the messages significantly impacted vaccination intent among the two groups. However, several messages significantly reduced intent to recommend vaccine to healthy adults among participants with tertiary education. The *DN(70%)* arm showed the highest drop in intent (−7.9 percentage points, 95% CI -12.6 to -3.2), followed by the *NF* arm (−5.7 percentage points, 95% CI -10.1 to -1.2), *DN* arm (−4.6 percentage points, 95% CI -8.8 to -0.3), and *RC(S)* arm (−4.5 percentage points, 95% CI -8.8 to -0.1). There were also significant reductions in intent between 5.5 to 6.6 percentage points among tertiary educated participants who were exposed to combination messages containing *PF* and *RC(S)*. However, participants without tertiary education had significantly lower intents to recommend elderly when exposed to combination messages containing *NF* (−7.2 percentage points, 95% CI -12.4 to -2.1) and *RC(SE)* (−5.5 percentage points, 95% CI -10.6 to -0.5). Apart from *DN(70%), HCW*, and combination messages containing *PF* and *RC(SE)*, all arms showed significant improvement in intent among those with tertiary education to recommend people with health conditions, ranging from 6.0 to 9.3 percentage points. The only exception was the *DN* arm whereby intent improved between 7.3 to 11.6 percentage points, irrespective of education level.

## Discussion

This study reports the results of one of the first experiments in the Southeast Asian region, and Malaysia specifically, that apply persuasive health messages to influence vaccine uptake and recommendation intentions. Hence, our results may serve as a reference benchmark of outcomes across various types of message frames from a middle-income country’s perspective.

Our estimates suggest that employed persuasive messages neither improved intent to vaccinate, nor recommendation intent to healthy and older individuals. In contrast, we find significant and sizeable effect of persuasive messages in improving the intent to recommend the vaccine to people with health conditions across several experimental arms.

Brief messages plus a short intervention time may have insufficiently addressed vaccine safety concerns to influence most of our outcomes. Moreover, consistent with the health belief model, concerns over vaccine safety are categorised as a perceived barrier towards vaccine uptake behaviour. Survey evidence among Malaysians documents both perceived benefits and perceived susceptibility as significant predictors of the intent to receive the COVID-19 vaccine in addition to barriers.[10] Hence, addressing only one predictor could be insufficient to influence vaccine hesitant individuals.

Attribute appeal is another possible reason driving the differences in recommendation intent between our studied outcomes. More than 80% of our participants agreed to recommend the vaccine to healthy adults at baseline. This observation suggests a general perception that healthy adults are fit enough to take the vaccine without any cause for safety concerns. Hence, addressing vaccine safety may not be a suitable nudge for the hesitant minority who may have deeper qualms about other issues. Conversely, about half of our participants were hesitant at baseline to recommend people with health issues. Such low baseline proportions may be driven by perceptions that vaccines are potentially harmful to individuals with pre-existing health conditions who may have higher susceptibility to vaccine adverse effects, given their poorer health state. This presumption is evidenced by the significant improvement in recommendation intent after exposure to several of our messages promoting vaccine safety, a key attribute that appealed to influenced participants. Similarly, vaccine safety may be a concern among participants who were hesitant to recommend the elderly, given that they are more frail and fragile to be recommended an intervention perceived as risky. However, the elderly are regarded highly in Asian societies such as Malaysia.[47] Given that our current sample skews towards younger individuals, recommending a perceived risky intervention to an elder may seem disrespectful. Therefore, persuasive vaccine safety messages proved insufficient to nudge those hesitant to recommend amidst an additional cultural barrier.

Our descriptive norm messages are grounded on the perceived sense of safety generated from knowledge that a vast majority are taking or have taken the COVID-19 vaccine, making it a social norm deemed as the right choice. However, this norms effect proved ineffective in significantly raising self-vaccination intent compared to the control message, consistent with COVID-19 vaccine studies involving norms.[22,23]. Helfinstein *et al*. also found that descriptive norms had little effect on risk recommendation to others, which reflects our negative observations with respect to vaccine recommendation.[48] However, we observe the *DN* message increasing recommendation intent to people with pre-existing health conditions. Message targeting may have made the *DN* message relatable to the recommended target group, since it highlights that many people with health conditions have tested and taken the COVID-19 vaccine.[49] However, the addition of *DN(70%)* weakened this effect. *DN(70%)* on its own reduced intentions to recommend healthy adults. This backfire could be due to perceptions that 70% of Malaysians, as stated in the *DN(70%)* message, is insufficient to be a convincing norm since mass media widely reports target inoculation rates of 80% by the government through the national immunisation programme.[50,51]

Both *NF* and *PF* messages induced opposite effects in two separate outcome measures. The *NF* message reduced intent to accept the COVID-19 vaccine, whilst the *PF* message improved intentions to recommend people with health conditions. The former result was similarly observed when the *DN(70%)* message was added, indicating that the descriptive norm message did not strengthen or dampen the effects from the negative attribute frame. Generally, studies have shown attribute frames to be more effective when framed positively rather than negatively.[52–55] However, Barnes & Colagiuri found that both positive and negative attribute framed messages increased intentions to accept a booster dose among COVID-19 vaccinated participants if the offered vaccine was unfamiliar or familiar respectively.[21] Their findings differed from our results possibly because our participants have not been vaccinated but were already familiar with the type of COVID-19 vaccine offered that was being widely promoted on mass media, given that our survey coincided with the national immunisation programme.[56–58] Inexperience towards the vaccine may have heightened negative safety perceptions arising from negative attribute framing whilst negating positive effects observed with positive attribute framing with respect to vaccination intent. A study involving influenza vaccine similarly found that participants who were exposed to negative framed messages had higher expectations or perceived severity of side effects as compared to their counterparts.[59] Interestingly, inexperience did not cloud positive perceptions arising from the *PF* message to drive improved intentions to recommend people with health conditions. Pre-conceived views that this target population is more susceptible to vaccine adverse effects given their poorer state of health may have been alleviated by this extra boost in safety perception.

Participants exposed to the *HCW* message did not show any significant changes in intent for all outcome measures. There are several possible reasons. The social norm cue used with reference to the majority of healthcare workers already vaccinated was probably ineffective due to participants being unable to identify with the reference population used.[23] Furthermore, the message may not have provided the personal touch and physical interaction from a healthcare provider necessary to invoke changes in intent, a condition which is observed among studies reporting raised vaccination intents [27,60,61]. This explanation is further supported by findings from Motta *et al*. whereby vaccination intent did not differ from the control group when the message encouraging vaccine uptake came from a medical expert.[62] Additionally, leveraging the Director General of Health’s influence, who is a government official, may portray him as accomplishing a bureaucratic task driven by political motives.[63] The use of a celebrity who is viewed as politically neutral yet popular, could prove more efficacious, as shown in a study which found celebrities inducing higher vaccine skepticism reductions compared to government officials or medical experts.[63] Interestingly, when both *HCW* and *DN(70%)* were combined, intent to recommend people with health conditions was significantly raised. This observation is probably borne from positive interactions between a low descriptive norm and a high injunctive norm. Recommendation from a convincing proportion of healthcare workers confers the perception that getting vaccinated is a socially desirable action that is expected to be done, which results in a high injunctive norm.[64] Habib *etal*. found that willingness to register as an organ donor increased when a low descriptive norm was combined with a high injunctive norm, as opposed to applying the norms individually.[65] This interaction arises by stoking a sense of responsibility to act after the incongruent norms makes salient regarding inconsistencies existing within the group. Although unmeasured, we believe this sense of responsibility to recommend was invoked from this similar interaction. Our finding thus expands knowledge on normative influence by proving such interactions also exist for behavior recommendation.

When risky choice framed messages were applied individually, only intent to recommend healthy adults was significantly negatively affected by *RC(S)*. The use of death rates from COVID-19 could be perceived as an irrelevant risk to healthy adults, since most deaths are associated with elderly and people with medical issues.[66] A mismatch with the target group for recommendation could have led to drops in intent. Moreover, the number of deaths featured on the message may not be convincing enough to require a need for healthy people to take the vaccine. However, based on shifts in the point estimate, this effect was slightly reduced when *DN(70%)* was added together, presumably because the higher dosage of pro-vaccination messages counteracted the negative effects of each message when applied individually.[31] A similar dose response interaction may be occurring when *DN(70%)* was combined with *RC(S*) to yield a significant increase in intent to recommend people with health conditions. Although *RC(S)* and *RC(SE)* addressed safety attributes which are relevant to elderly and people with health conditions, their effects did not differ from the control message when applied alone. A possible reason lies with the message bringing attention to possible health risks associated with the vaccine such as deaths or blood clots. Despite the probability favouring vaccine uptake, the mention of these health risks may have caused hesitant individuals to remain hesitant for fear of recommending something harmful.

Our analysis on heterogeneity treatment effects revealed varied impacts of different messages for each socio-demographic variable. Intent to vaccinate for both older participants and males were negatively influenced by a negative attribute frame. Studies show older people have higher risk perceptions towards health related risks.[67] This characteristic makes them more susceptible to negatively framed attribute messages as negative frames heighten risk perception. Studies have also shown that men tend to be more optimistic about perceived susceptibility and severity from COVID-19,[68,69] rendering males being more likely to take a risk with contracting the virus as compared to taking a vaccine that is perceived unsafe due to the negative attribute framing effect. Our findings highlight the damaging effect such frames can cause amongst males who generally have higher vaccination intentions compared to females.[70]

Most of the messages which induced positive intents to recommend people with health conditions impacted the older age group, males, and those with a tertiary education. There are several postulations to this pattern of results. Studies show that self-esteem increases with age.[71,72] This may confer older people with more confidence to recommend the vaccine if there is information that supports this action. Moreover, our youths may be more hesitant to recommend even when nudged as Malaysia practices a collectivist culture.[47] People with medical condition tend to be older, which makes it more challenging for youths to recommend due to social hierarchy barriers. In terms of males having higher intents, we postulate differences in risk acceptance as a possible explanation. Recommending a health intervention involves some risk taking since it advocates something that may bring risk to another individual. Studies have shown males have higher risk taking behaviour compared to females.[73] Therefore, males could be more willing to accept risks associated with recommendation, thus responding positively to more of the messages compared to females. On the other hand, behavioural differences to recommend based on education level are probably related to cognitive capabilities to synthesize information and perceived vaccine safety. People with tertiary education could have understood and synthesised the health messages better to infer that the vaccine was safe to be used by people with health issues. Being higher educated also gives more confidence and a higher sense of social responsibility to recommend.

Individuals with tertiary education were also more impacted by messages which reduced intent to recommend healthy adults. A deeper synthesis of messages by those who have higher education does not necessarily produce positive results and could backfire instead. These people may tend to have more complex interpretations amidst wider information obtained from various sources, resulting in certain messages inducing negative responses. Studies have shown that there is strong associations between education level and extent of COVID-19 related knowledge, both factual and perceived.[74,75] Coupled with a lesser perceived severity of the virus by more educated individuals, these messages may have been interpreted with a risk benefit analysis to suggest healthy individuals not requiring the vaccine.[75]

### Limitations

Our experiment exhibits the following limitations. Study outcomes measured how messages affect intent and do not really indicate whether participants would actually receive or recommend the vaccine in reality. Although actual vaccination behaviour should be the prime outcome of interest, intent has been shown to be a strong predictor for behavioural actions over various contexts, even for actual vaccination uptake.[76] However, significant intention-behavior gaps for vaccination has been shown to exist,[77] with a study even concluding that nudges are ineffective at significantly raising actual COVID-19 vaccination rates.[18] Previous research have also shown differing results when applying behavioral nudges to promote COVID-19 vaccination under experimental conditions versus in the field.[78] These findings underscore the need to field test behavioural interventions that are proven successful in survey experiments to confirm their true effectiveness under real world conditions.

The extent of misinformation that participants were exposed to prior to our experiment was not measured. Misinformation has been proven to significantly affect vaccination intent.[43] Actual vaccination rates declined depending on the theme and quantity of misinformation exposure.[13] Therefore, misinformation exposure may be a strong predictor for resisting nudges from health messages. Future studies should find ways of incorporating this measure to further elucidate true effectiveness of messages under various levels of misinformation exposure.

The dynamic nature of the COVID-19 pandemic may have altered attitudes towards the COVID-19 vaccines since our experiment was initiated. This is especially so after the vaccines have been safely rolled out and shown to be effective as time progresses. Hence, the efficacy of these messages may have changed over the course of the pandemic.

Lastly, we did not specify any particular COVID-19 vaccine when asking participants to take or recommend. During the experimental survey roll out, vaccines from three different companies were widely mentioned in Malaysia, namely Pfizer-BioNTech, Oxford/Astra Zeneca, and Sinovac.[58,79] Each of these vaccines were developed using different technologies to yield differing effectiveness and safety profiles. The public may hold differing views about the vaccines based on the familiarity of the technology used to develop them. Hence, we were unsure whether responses obtained were based on a particular vaccine in mind or aggregated in nature.

### Further work

Explanations regarding behavioural responses observed were inferred based on past research. More in depth qualitative research based on theoretical frameworks should be conducted to gain a firmer understanding on how these messages affect individual perceptions that result in provided responses. Additionally, more research should be conducted to understand the science behind individuals recommending healthcare interventions to others, as this aspect of knowledge in the health behavioural field is scarce.

## Conclusion

Despite safety being the main concern for COVID-19 vaccine hesitancy, crafting messages that focus solely on this attribute does not significantly improve vaccination intent or vaccine recommendation, except to people who have pre-existing health conditions. Our findings prove that addressing a single attribute that is of highest concern for vaccine hesitancy may not necessarily improve vaccination uptake

We documented several examples where combining messages weakened or strengthened intent, thus providing further proof about message interactions between different frames. A deeper understanding of such interactions is needed, especially when conducting health promotion campaigns that utilise a series of messages together to influence individual decision making.

Our study suggests two important findings. Firstly, persuasive messages aimed at influencing vaccination decisions should incorporate a combination of factors linked to hesitancy instead of only one single attribute. Lastly, messages incorporating safety dimensions are capable of updating the belief of individuals to advocate an intervention that was previously deemed risky to a vulnerable population. This evidence suggests that persuasive messages should emphasize on safety when promoting recommendation of novel health interventions to individuals with pre-existing health conditions, especially if the intervention is perceived as harmful to them.

## Data Availability

The dataset used for this study belongs to the Ministry of Health, Malaysia. Hence, the dataset may be available from the corresponding author via a formal request through relevant authorities at the Ministry of Health, Malaysia.

## Informed Consent Statement

Informed consent was obtained from all subjects involved in the study. Participants consented to participation if they clicked on an informed consent button after reading the online survey’s introductory section and participant information sheet.

## Ethics statements

### Patient consent for publication

Not required.

### Patient and Public Involvement statement

Patients or the public were not involved in the design, or conduct, or reporting, or dissemination plans of our research.

### Ethics approval

Ethical approval for this study was granted by the Medical Research Ethics Committee of the Ministry of Health Malaysia on 25 Feb 2021 (KKM/NIHSEC/ P21-130 (4)).

## Acknowledgments

We would like to thank the Director General of Health Malaysia for his permission to publish this article. We would also like to thank the team at Dynata for their efficient service in making the data collection process for this study a success.

## Contributors

NYLH, YLW, YKL, NML and JCF contributed to the conception and design of the study. NYLH, YLW, YKL, NML, JKH, EW, KP, NHAS and AI contributed to content and design of experimented messages, and questionnaire development. Questionnaire and message validation was conducted by NYLH, YLW, YKL, NML, KP and NHAS. Project management was handled by NYLH. NYLH and CTL conducted statistical analysis. JCF was consulted for data analysis. Visualisations of results were prepared by NYHL. NYHL wrote the original draft of the manuscript. JCF supervised the drafting of the manuscript. All authors interpreted the results and critically reviewed the drafts of this manuscript. All authors read and approved the final manuscript.

## Exclusive License statement

I, the Submitting Author has the right to grant and does grant on behalf of all authors of the Work (as defined in the below author licence), an exclusive licence and/or a non-exclusive licence for contributions from authors who are: i) UK Crown employees; ii) where BMJ has agreed a CC-BY licence shall apply, and/or iii) in accordance with the terms applicable for US Federal Government officers or employees acting as part of their official duties; on a worldwide, perpetual, irrevocable, royalty-free basis to BMJ Publishing Group Ltd (“BMJ”) its licensees and where the relevant Journal is co-owned by BMJ to the co-owners of the Journal, to publish the Work in BMJ Global Health and any other BMJ products and to exploit all rights, as set out in our license.

The Submitting Author accepts and understands that any supply made under these terms is made by BMJ to the Submitting Author unless you are acting as an employee on behalf of your employer or a postgraduate student of an affiliated institution which is paying any applicable article publishing charge (“APC”) for Open Access articles. Where the Submitting Author wishes to make the Work available on an Open Access basis (and intends to pay the relevant APC), the terms of reuse of such Open Access shall be governed by a Creative Commons licence – details of these licences and which Creative Commons licence will apply to this Work are set out in our licence referred to above.

## Author Reflexivity Statement

### 1. How does this study address local research and policy priorities?

This study addresses the need to investigate what message frames are effective at influencing Malaysians to take up the COVID-19 vaccine, as well as recommending it to others in society. At the time of the study, Malaysia was just only rolling out the COVID-19 vaccination programme and there was an urgent need to determine what message frames would be effective to bolster vaccination registration and uptake rates.

### 2. How were local researchers involved in study design?

The first group of local researchers who initiated the research question and idea were NYLH and YLW. Both researchers were based at the Institute for Clinical Research at the Malaysian National Institute of Health and were well connected and capable of conducting, leading, and organising local and international research collaborations. The second group of researchers who were invited by the core team to plan and discuss the conduct of the research project were local researchers based in local academic, research and government policy institutions (YKL, NML, KP, NHAS and AI). These researchers were from a diverse background and had intermediate to advanced research skills that helped solidify the study design. The third group of local researchers consisted were from a non-governmental organisation (JKH and EW) and had specific experience in risk communication, which is a vital part of this study. JCF was the only research member who was based abroad in a high-income country. JCF was invited to join the study to provide his expertise related to behavioral economics. Therefore, almost the entire research team who are based in a middle-income country were actively involved with the study design. Our researcher that is based in a high income country was mainly responsible for providing guidance and support in terms of ensuring study design and interventions used were valid and scientifically sound.

### 3. How has funding been used to support the local research team?

Funding was mainly used to engage the services of an international market research compan (i.e. Dynata) to conduct the study’s online survey through their survey panel in Malaysia.

### 4. How are research staff who conducted data collection acknowledged?

All members of the research team were included into the authorship of this paper as a form of acknowledgement for their contributions offered.

### 5. Do all members of the research partnership have access to study data?

All members of the partnership have access to the data except JCF. This exception is due to data confidentiality and security restrictions imposed by the Malaysian government for government owned data. Data cannot be transferred abroad unless a formal application is applied by the foreign party.

### 6. How was data used to develop analytical skills within the partnership?

JCF was consulted by members of the Malaysian research team who were tasked with data analysis. Knowledge transfer obtained from consultations provided sufficient analytical skills needed to analyse data.

### 7. How have research partners collaborated in interpreting study data?

Two online meetings were held among all study team members during the process of study data interpretations. Meetings involved presenting summary of data collected, discussion of analysis plans, presentation of draft and finalized results, and data interpretations. Three other separate online meetings were held between NYLH, CTL and JCF to discuss further queries and data interpretation during manuscript write up.

### 8. How were research partners supported to develop writing skills?

NYLH, who is the main author of the current manuscript, was guided and supported by JCF who is a senior academic at the London School of Economics and Political Science. Guidance and support entailed writing style, techniques to formulate critical discussions, and assistance in editing the final manuscript.

### 9. How will research products be shared to address local needs?

Research outputs were shared to local and international stakeholders who were involved with risk communication activities to improve COVID-19 vaccination uptake among Malaysians. These included the Health Education Division at the Malaysian Ministry of Health, the World Health Organisation (Western Pacific Region) and UNICEF, Malaysia.

### 10. How is the leadership, contribution and ownership of this work by LMIC researchers recognised within the authorship?

Authors NYLH and JCF worked as part of the authorship team in developing this manuscript. Their contribution has been recognised as joint first and joint last authors respectively. Hence both middle income and high income country authors share main authorships for this paper, amidst an authorship team that is predominantly based in a middle-income country.

### 11. How have early career researchers across the partnership been included within the authorship team?

We have included an early career researcher (NML) within the authorship team. She attended all project meetings, contributed to the literature review, and assisted with both the development and validation of the survey questionnaire and experimental messages. We acknowledge that she is based in a middle-income country.

### 12. How has gender balance been addressed within the authorship?

Seven authors are male (NYLH, YLW, CTL, YKL, JKH, AI and JCF) and AI) and four authors are female (NML, EW, KP and NHAS). We admit that gender balance was slightly skewed towards males in this study’s authorship list. We hope to ensure a more gender balanced group of authors in the future.

### 13. How has the project contributed to training of LMIC researchers?

The project has exposed and taught Malaysian researchers in the team on how to conduct behavioral insights research work, along with data analysis techniques arising from such projects.

### 14. How has the project contributed to improvements in local infrastructure?

This project has not directly contributed to improvements in local infrastructure.

### 15. What safeguarding procedures were used to protect local study participants and researchers?

Local study participants were safeguarded by not collecting their personal identifiers throughout the online survey. Dynata does not share personal information of participants who responded to the survey, in accordance to data privacy policies. This question is not directly applicable to researchers as the study conduct only requires recruited participants to answer an online survey.

