## Supplementary material for "When do persuasive messages on vaccine safety steer COVID-19 vaccine acceptance and recommendations? Behavioral insights from a randomised controlled experiment in Malaysia": online supplemental material

### Methods

### Study setting during recruitment period

Malaysia was experiencing a surge of infections in April 2021, with over 3,300 daily cases and almost 1,500 total deaths reported at the start of our experiment.[1] By the end of our experiment, daily cases steadily increased to reach a peak of over 7,700 cases, with cumulative deaths standing at 3,378.[2] Malaysia's COVID-19 immunisation programme was initiated at the end of February 2021.[3] Our experiment coincided closely with the second and third phase of the programme which began in April and May 2021 respectively. These two phases were targeted at the general adult population.

### Stratified sampling

Malaysia is composed of several major ethnicities. Bumiputera, which consist of Malays and the indigenous people of Malaysia, accounted for about 70% of the population.[4] This is followed by Chinese (≈23%) and Indians (≈7%). The sex ratio among Malaysian citizens stands at 102 males per 100 females. There is a sizable proportion of young Malaysian in the country, with approximately 53% of the total adult population aged between 18 to 39 years. Middle age (40 to 59 years old) and the elderly accounted for approximately 31% and 16% of the population respectively. In terms of household income, Malaysia categorizes citizens into three distinct groups; Bottom 40% (B40), Middle 40% (M40) and Top 20% (T20).[5] These categorisations represent percentages of the country's population in terms of household income ranging from the bottom 40% to the top 20%. Except for age, stratified recruitment was conducted according to approximate national ratios for gender, ethnicity and household income. Due to our survey panel's limitation to sample for older participants, we inflated and deflated the target sampling proportion for the younger and older age group by about 10% and 12% respectively.

**Message design**

Messages were designed with a standardised dimension of 1080 x 1350 pixels in order to look similar with messages commonly found on social media posts and is conveniently displayed on computer monitors or smartphones. Font sizes used for all messages were standardised. Numbers or words which indicated a numerical or statistical meaning were printed using yellow colour fonts that were slightly enlarged to draw extra attention. The last sentence in the rally slogan; "It's safe and effective!", was printed in a green font to psychologically invoke feelings of safety about the vaccine.[6]
