## supplemental table S1 for "When do persuasive messages on vaccine safety steer COVID-19 vaccine acceptance and recommendations? Behavioral insights from a randomised controlled experiment in Malaysia"

**Table S1: Baseline characteristics of survey participants stratified according to experimental arms**

| | | DN (70%) | DN | HCW | NF | PF | RC(S) | RC(SE) | Control | DN(70%) + DN | DN(70%) + HCW | DN(70%) + NF | DN(70%) + PF | DN(70%) + RC(S) | DN(70%) + RC(SE) | T-test/Chi square |
|---|---|---|---|---|---|---|---|---|---|---|---|---|---|---|---|---|
| | | Mean±SD or N(%) | Mean±SD or N(%) | Mean±SD or N(%) | Mean±SD or N(%) | Mean±SD or N(%) | Mean±SD or N(%) | Mean±SD or N(%) | Mean±SD or N(%) | Mean±SD or N(%) | Mean±SD or N(%) | Mean±SD or N(%) | Mean±SD or N(%) | Mean±SD or N(%) | Mean±SD or N(%) | P-value |
| Age | | 37±11.6 | 36±11.9 | 36±11.8 | 36±11.5 | 36±11.9 | 35±11.3 | 36±11.7 | 36±11.6 | 36±11.7 | 36±11.9 | 36±11.8 | 36±11.4 | 36±11.9 | 36±11.6 | 0.999 |
| Sex | Male | 210 (50.6) | 203 (49.4) | 206 (50.2) | 206 (49.8) | 208 (50.2) | 210 (50.5) | 205 (50.0) | 207 (50.5) | 211 (50.8) | 207 (49.8) | 207 (50.1) | 211 (50.8) | 211 (50.8) | 205 (50.0) | 1 |
| Education level | Tertiary education | 194(46.7) | 218(53.0) | 205(50.0) | 209(50.5) | 182(44.0) | 198(47.6) | 200(48.8) | 217(52.9) | 194(46.7) | 225(54.1) | 209(50.6) | 204(49.2) | 215(51.8) | 195(47.6) | 0.169 |
| Intent to vaccinate | Definitely not | 5(1.2) | 8(1.9) | 8(2.0) | 9(2.2) | 9(2.2) | 7(1.7) | 8(2.0) | 5(1.2) | 7(1.7) | 9(2.2) | 7(1.7) | 9(2.2) | 11(2.7) | 4(1.0) | 0.968 |
| | Probably not | 32(7.7) | 28(6.8) | 25(6.1) | 28(6.8) | 26(6.3) | 29(7.0) | 30(7.3) | 32(7.8) | 34(8.2) | 33(7.9) | 20(4.8) | 29(7.0) | 27(6.5) | 17(4.1) | |
| | Probably yes | 123(29.6) | 118(28.7) | 119(29.0) | 120(29.0) | 133(32.1) | 133(32.0) | 121(29.5) | 111(27.1) | 129(31.1) | 122(29.3) | 116(28.1) | 132(31.8) | 123(29.6) | 124(30.2) | |
| | Definitely yes | 255(61.4) | 257(62.5) | 258(62.9) | 257(62.1) | 246(59.4) | 247(59.4) | 251(61.2) | 262(63.9) | 245(59.0) | 252(60.6) | 270(65.4) | 245(59.0) | 254(61.2) | 265(64.6) | |
| Intent to recommend: | | | | | | | | | | | | | | | | |
| Healthy adults | Strongly disagree | 8(1.9) | 8(1.9) | 5(1.2) | 7(1.7) | 6(1.4) | 7(1.7) | 7(1.7) | 6(1.5) | 7(1.7) | 5(1.2) | 8(1.9) | 5(1.2) | 5(1.2) | 3(0.7) | 0.92 |
| | Disagree | 15(3.6) | 11(2.7) | 11(2.7) | 9(2.2) | 13(3.1) | 10(2.4) | 18(4.4) | 8(2.0) | 12(2.9) | 22(5.3) | 13(3.1) | 17(4.1) | 17(4.1) | 9(2.2) | |
| | Not sure | 46(11.1) | 42(10.2) | 43(10.5) | 53(12.8) | 49(11.8) | 37(8.9) | 43(10.5) | 36(8.8) | 48(11.6) | 40(9.6) | 37(9.0) | 49(11.8) | 42(10.1) | 40(9.8) | |
| | Agree | 198(47.7) | 202(49.1) | 183(44.6) | 198(47.8) | 204(49.3) | 210(50.5) | 195(47.6) | 206(50.2) | 204(49.2) | 214(51.4) | 213(51.6) | 201(48.4) | 196(47.2) | 209(51.0) | |
| | Strongly agree | 148(35.7) | 148(36.0) | 168(41.0) | 147(35.5) | 142(34.3) | 152(36.5) | 147(35.9) | 154(37.6) | 144(34.7) | 135(32.5) | 142(34.4) | 143(34.5) | 155(37.3) | 149(36.3) | |
| Elderly | Strongly disagree | 11(2.7) | 10(2.4) | 12(2.9) | 11(2.7) | 9(2.2) | 11(2.6) | 13(3.2) | 13(3.2) | 11(2.7) | 18(4.3) | 8(1.9) | 13(3.1) | 19(4.6) | 9(2.2) | 0.622 |
| | Disagree | 36(8.7) | 28(6.8) | 23(5.6) | 27(6.5) | 28(6.8) | 28(6.7) | 30(7.3) | 25(6.1) | 26(6.3) | 35(8.4) | 16(3.9) | 24(5.8) | 20(4.8) | 26(6.3) | |
| | Not sure | 68(16.4) | 75(18.2) | 69(16.8) | 78(18.8) | 79(19.1) | 76(18.3) | 73(17.8) | 64(15.6) | 96(23.1) | 80(19.2) | 72(17.4) | 79(19.0) | 61(14.7) | 70(17.1) | |
| | Agree | 161(38.8) | 164(39.9) | 153(37.3) | 156(37.7) | 165(39.9) | 169(40.6) | 156(38.0) | 182(44.4) | 159(38.3) | 156(37.5) | 173(41.9) | 165(39.8) | 177(42.7) | 167(40.7) | |
| | Strongly agree | 139(33.5) | 134(32.6) | 153(37.3) | 142(34.3) | 133(32.1) | 132(31.7) | 138(33.7) | 126(30.7) | 123(29.6) | 127(30.5) | 144(34.9) | 134(32.3) | 138(33.3) | 138(33.7) | |
| People with health conditions | Strongly disagree | 15(3.6) | 19(4.6) | 20(4.9) | 22(5.3) | 18(4.3) | 26(6.3) | 24(5.9) | 21(5.1) | 18(4.3) | 24(5.8) | 19(4.6) | 24(5.8) | 26(6.3) | 16(3.9) | 0.941 |
| | Disagree | 63(15.2) | 59(14.4) | 40(9.8) | 58(14.0) | 55(13.3) | 56(13.5) | 60(14.6) | 49(12.0) | 58(14.0) | 60(14.4) | 49(11.9) | 62(14.9) | 46(11.1) | 51(12.4) | |
| | Not sure | 128(30.8) | 113(27.5) | 121(29.5) | 126(30.4) | 133(32.1) | 128(30.8) | 119(29.0) | 120(29.3) | 131(31.6) | 121(29.1) | 120(29.1) | 137(33.0) | 124(29.9) | 131(32.0) | |
| | Agree | 114(27.5) | 120(29.2) | 136(33.2) | 110(26.6) | 113(27.3) | 103(24.8) | 115(28.0) | 128(31.2) | 125(30.1) | 123(29.6) | 124(30.0) | 110(26.5) | 126(30.4) | 124(30.2) | |
| | Strongly agree | 95(22.9) | 100(24.3) | 93(22.7) | 98(23.7) | 95(22.9) | 103(24.8) | 92(22.4) | 92(22.4) | 83(20.0) | 88(21.2) | 101(24.5) | 82(19.8) | 93(22.4) | 88(21.5) | |
| Negative vaccine attitude | No | 272(65.5) | 275(66.9) | 266(64.9) | 278(67.1) | 267(64.5) | 256(61.5) | 261(63.7) | 267(65.1) | 273(65.8) | 261(62.7) | 281(68.0) | 253(61.0) | 266(64.1) | 277(67.6) | 0.603 |
