## supplemental table S2 for "When do persuasive messages on vaccine safety steer COVID-19 vaccine acceptance and recommendations? Behavioral insights from a randomised controlled experiment in Malaysia"

**Table S2: Average marginal effects for intention to accept the COVID-19 vaccine in each experimental arm relative to control arm**

| | **Intention to vaccinate**<br>Marginal effects<br>[95% Confidence Interval] |
|---|---|
| **DN(70%)** | |
| Definitely no | 0.00392<br>[-0.000780,0.00861] |
| Probably no | 0.00516<br>[-0.00106,0.0114] |
| Probably yes | 0.0163<br>[-0.00324,0.0359] |
| Definitely yes | -0.0254<br>[-0.0557,0.00484] |
| **DN** | |
| Definitely no | -0.000924<br>[-0.00519,0.00334] |
| Probably no | -0.00133<br>[-0.00745,0.00480] |
| Probably yes | -0.00446<br>[-0.0250,0.0161] |
| Definitely yes | 0.00671<br>[-0.0242,0.0376] |
| **HCW** | |
| Definitely no | 0.000472<br>[-0.00394,0.00488] |
| Probably no | 0.000659<br>[-0.00550,0.00682] |
| Probably yes | 0.00217<br>[-0.0182,0.0225] |
| Definitely yes | -0.00330<br>[-0.0342,0.0276] |
| **NF** | |
| Definitely no | 0.00519*<br>[0.000312,0.0101] |
| Probably no | 0.00671*<br>[0.000347,0.0131] |
| Probably yes | 0.0210*<br>[0.00134,0.0406] |
| Definitely yes | -0.0329*<br>[-0.0633,-0.00240] |
| **PF** | |
| Definitely no | -0.000954<br>[-0.00517,0.00326] |
| Probably no | -0.00137<br>[-0.00742,0.00468] |

| | |
|---|---|
| Probably yes | -0.00461<br>[-0.0249,0.0157] |
| Definitely yes | 0.00693<br>[-0.0236,0.0375] |
| **RC(S)** | |
| Definitely no | 0.000399<br>[-0.00393,0.00473] |
| Probably no | 0.000558<br>[-0.00550,0.00661] |
| Probably yes | 0.00184<br>[-0.0182,0.0219] |
| Definitely yes | -0.00280<br>[-0.0332,0.0276] |
| **RC(SE)** | |
| Definitely no | 0.00139<br>[-0.00307,0.00585] |
| Probably no | 0.00191<br>[-0.00422,0.00804] |
| Probably yes | 0.00623<br>[-0.0138,0.0262] |
| Definitely yes | -0.00954<br>[-0.0401,0.0210] |
| **DN(70%)+DN** | |
| Definitely no | 0.00161<br>[-0.00281,0.00603] |
| Probably no | 0.00220<br>[-0.00385,0.00825] |
| Probably yes | 0.00716<br>[-0.0126,0.0269] |
| Definitely yes | -0.0110<br>[-0.0411,0.0192] |
| **DN(70%)+HCW** | |
| Definitely no | 0.00158<br>[-0.00288,0.00603] |
| Probably no | 0.00216<br>[-0.00395,0.00826] |
| Probably yes | 0.00703<br>[-0.0128,0.0269] |
| Definitely yes | -0.0108<br>[-0.0412,0.0196] |
| **DN(70%)+NF** | |
| Definitely no | 0.00565*<br>[0.000671,0.0106] |
| Probably no | 0.00726*<br>[0.000827,0.0137] |

| | |
|---|---|
| Probably yes | 0.0226* |
| | [0.00288,0.0423] |
| Definitely yes | -0.0355* |
| | [-0.0661,-0.00485] |
| **DN(70%)+PF** | |
| Definitely no | 0.00241 |
| | [-0.00211,0.00693] |
| Probably no | 0.00325 |
| | [-0.00286,0.00936] |
| Probably yes | 0.0105 |
| | [-0.00922,0.0302] |
| Definitely yes | -0.0161 |
| | [-0.0464,0.0141] |
| **DN(70%)+RC(S)** | |
| Definitely no | 0.000300 |
| | [-0.00407,0.00467] |
| Probably no | 0.000420 |
| | [-0.00571,0.00655] |
| Probably yes | 0.00139 |
| | [-0.0189,0.0217] |
| Definitely yes | -0.00211 |
| | [-0.0329,0.0287] |
| **DN(70%)+RC(SE)** | |
| Definitely no | 0.00267 |
| | [-0.00198,0.00733] |
| Probably no | 0.00359 |
| | [-0.00265,0.00983] |
| Probably yes | 0.0115 |
| | [-0.00850,0.0316] |
| Definitely yes | -0.0178 |
| | [-0.0486,0.0130] |
| *N* | 5784 |

95% confidence intervals in brackets
* $p < 0.05$, ** $p < 0.01$, *** $p < 0.001$
