## supplemental table S3 for "When do persuasive messages on vaccine safety steer COVID-19 vaccine acceptance and recommendations? Behavioral insights from a randomised controlled experiment in Malaysia"

**Table S3: Average marginal effects for intention to recommend the COVID-19 vaccine to healthy adults, elderly, and people with any pre-existing health conditions, in each experimental arm relative to control arm.**

| | **Healthy adults** Marginal effects [95% Confidence Interval] | **Elderly** Marginal effects [95% Confidence Interval] | **Health condition** Marginal effects [95% Confidence Interval] |
|---|---|---|---|
| **DN(70%)** | | | |
| Disagree | 0.0148$^{*}$ [0.00255,0.0271] | 0.0121 [-0.00394,0.0281] | -0.00964 [-0.0298,0.0105] |
| Not sure | 0.0237$^{*}$ [0.00411,0.0432] | 0.0148 [-0.00480,0.0344] | -0.00919 [-0.0284,0.0100] |
| Agree | -0.0385$^{*}$ [-0.0702,-0.00673] | -0.0269 [-0.0625,0.00873] | 0.0188 [-0.0204,0.0581] |
| **DN** | | | |
| Disagree | 0.0111 [-0.00108,0.0232] | -0.000137 [-0.0160,0.0157] | -0.0411$^{***}$ [-0.0616,-0.0205] |
| Not sure | 0.0174 [-0.00167,0.0365] | -0.000165 [-0.0193,0.0189] | -0.0391$^{***}$ [-0.0588,-0.0195] |
| Agree | -0.0285 [-0.0596,0.00271] | 0.000302 [-0.0347,0.0353] | 0.0802$^{***}$ [0.0405,0.120] |
| **HCW** | | | |
| Disagree | 0.00320 [-0.00891,0.0153] | 0.0106 [-0.00536,0.0266] | -0.0150 [-0.0352,0.00529] |
| Not sure | 0.00485 [-0.0135,0.0232] | 0.0130 [-0.00653,0.0326] | -0.0143 [-0.0337,0.00507] |
| Agree | -0.00806 [-0.0385,0.0224] | -0.0236 [-0.0592,0.0119] | 0.0293 [-0.0103,0.0688] |
| **NF** | | | |
| Disagree | 0.0100 [-0.00207,0.0221] | 0.00806 [-0.00783,0.0240] | -0.0187 [-0.0387,0.00132] |
| Not sure | 0.0157 [-0.00323,0.0347] | 0.00984 [-0.00954,0.0292] | -0.0179 [-0.0370,0.00128] |
| Agree | -0.0258 [-0.0568,0.00527] | -0.0179 [-0.0532,0.0174] | 0.0366 [-0.00246,0.0756] |
| **PF** | | | |
| Disagree | 0.0102 [-0.00189,0.0222] | 0.00514 [-0.0105,0.0208] | -0.0288$^{**}$ [-0.0489,-0.00859] |
| Not sure | 0.0159 [-0.00293,0.0348] | 0.00624 [-0.0128,0.0253] | -0.0275$^{**}$ [-0.0469,-0.00821] |
| Agree | -0.0261 [-0.0569,0.00479] | -0.0114 [-0.0461,0.0233] | 0.0563$^{**}$ [0.0171,0.0955] |
| **RC(S)** | | | |
| Disagree | 0.0163$^{**}$ | 0.0106 | -0.0171 |

| | | | |
|---|---|---|---|
| | [0.00393,0.0287] | [-0.00526,0.0265] | [-0.0372,0.00287] |
| Not sure | 0.0262** | 0.0130 | -0.0164 |
| | [0.00637,0.0461] | [-0.00641,0.0324] | [-0.0356,0.00278] |
| Agree | -0.0425** | -0.0236 | 0.0336 |
| | [-0.0746,-0.0104] | [-0.0588,0.0117] | [-0.00553,0.0726] |
| **RC(SE)** | | | |
| Disagree | 0.00534 | 0.00154 | -0.0148 |
| | [-0.00663,0.0173] | [-0.0142,0.0173] | [-0.0349,0.00533] |
| Not sure | 0.00818 | 0.00186 | -0.0141 |
| | [-0.0101,0.0265] | [-0.0172,0.0209] | [-0.0334,0.00513] |
| Agree | -0.0135 | -0.00339 | 0.0289 |
| | [-0.0438,0.0168] | [-0.0382,0.0314] | [-0.0104,0.0682] |
| **DN(70%)+DN** | | | |
| Disagree | 0.0127* | 0.00193 | -0.0213* |
| | [0.000460,0.0249] | [-0.0135,0.0174] | [-0.0415,-0.00114] |
| Not sure | 0.0201* | 0.00234 | -0.0204* |
| | [0.000739,0.0394] | [-0.0163,0.0210] | [-0.0398,-0.00107] |
| Agree | -0.0328* | -0.00427 | 0.0417* |
| | [-0.0643,-0.00125] | [-0.0383,0.0298] | [0.00238,0.0811] |
| **DN(70%)+HCW** | | | |
| Disagree | 0.00418 | -0.00548 | -0.0240* |
| | [-0.00782,0.0162] | [-0.0209,0.00989] | [-0.0441,-0.00396] |
| Not sure | 0.00637 | -0.00653 | -0.0230* |
| | [-0.0119,0.0246] | [-0.0248,0.0118] | [-0.0423,-0.00372] |
| Agree | -0.0105 | 0.0120 | 0.0470* |
| | [-0.0408,0.0197] | [-0.0217,0.0457] | [0.00790,0.0862] |
| **DN(70%)+NF** | | | |
| Disagree | 0.00936 | 0.0141 | -0.0123 |
| | [-0.00304,0.0218] | [-0.00210,0.0302] | [-0.0326,0.00789] |
| Not sure | 0.0146 | 0.0173 | -0.0118 |
| | [-0.00477,0.0340] | [-0.00254,0.0371] | [-0.0311,0.00755] |
| Agree | -0.0240 | -0.0313 | 0.0241 |
| | [-0.0558,0.00778] | [-0.0673,0.00462] | [-0.0154,0.0637] |
| **DN(70%)+PF** | | | |
| Disagree | 0.0101 | 0.0159 | -0.0198 |
| | [-0.00188,0.0221] | [-0.0000648,0.0319] | [-0.0398,0.000177] |
| Not sure | 0.0159 | 0.0196 | -0.0190 |
| | [-0.00292,0.0347] | [-0.0000347,0.0392] | [-0.0381,0.000176] |
| Agree | -0.0260 | -0.0355 | 0.0387 |
| | [-0.0568,0.00477] | [-0.0711,0.0000810] | [-0.000198,0.0777] |
| **DN(70%)+RC(S)** | | | |
| Disagree | 0.0124* | 0.00325 | -0.0235* |

| | | | |
|---|---|---|---|
| | [0.000140,0.0247] | [-0.0129,0.0194] | [-0.0436,-0.00327] |
| Not sure | 0.0196* | 0.00393 | -0.0225* |
| | [0.000215,0.0390] | [-0.0156,0.0235] | [-0.0418,-0.00310] |
| Agree | -0.0320* | -0.00718 | 0.0459* |
| | [-0.0636,-0.000404] | [-0.0428,0.0285] | [0.00658,0.0853] |
| **DN(70%)+RC(SE)** | | | |
| Disagree | 0.00744 | 0.0136 | -0.00524 |
| | [-0.00497,0.0199] | [-0.00261,0.0299] | [-0.0254,0.0149] |
| Not sure | 0.0115 | 0.0167 | -0.00499 |
| | [-0.00770,0.0307] | [-0.00317,0.0366] | [-0.0242,0.0142] |
| Agree | -0.0190 | -0.0304 | 0.0102 |
| | [-0.0506,0.0127] | [-0.0665,0.00577] | [-0.0291,0.0496] |
| *N* | 5784 | 5784 | 5784 |

95% confidence intervals in brackets
$^{*}$ $p < 0.05$, $^{**}$ $p < 0.01$, $^{***}$ $p < 0.001$
