## supplemental table S4 for "When do persuasive messages on vaccine safety steer COVID-19 vaccine acceptance and recommendations? Behavioral insights from a randomised controlled experiment in Malaysia"

**Table S4: Chi-square analysis describing associations between all experimental arms and proportion of hesitant participants who cited reasons of vaccine safety or side effect concerns after message exposure.**

| Hesitancy to: | Intention to vaccinate | | Recommend healthy adults | | Recommend elderly | | Recommend people with health conditions | |
|---|---|---|---|---|---|---|---|---|
| Worried about the safety or side effects of the vaccine. | No (%) | Yes (%) | No (%) | Yes (%) | No (%) | Yes (%) | No (%) | Yes (%) |
| DN(70%) | 22.78 | 77.22 | 30.00 | 70.00 | 20.00 | 80.00 | 18.95 | 81.05 |
| DN | 27.74 | 72.26 | 28.81 | 71.19 | 20.43 | 79.57 | 20.92 | 79.08 |
| HCW | 22.86 | 77.14 | 33.33 | 66.67 | 14.85 | 85.15 | 14.55 | 85.45 |
| NF | 32.67 | 67.33 | 34.48 | 65.52 | 22.86 | 77.14 | 25.53 | 74.47 |
| PF | 21.92 | 78.08 | 31.15 | 68.85 | 14.15 | 85.85 | 20.24 | 79.76 |
| RC(S) | 26.67 | 73.33 | 29.51 | 70.49 | 18.18 | 81.82 | 19.78 | 80.22 |
| RC(SE) | 20.53 | 79.47 | 19.3 | 80.7 | 25.24 | 74.76 | 19.34 | 80.66 |
| Control | 18.71 | 81.29 | 38.46 | 61.54 | 23.08 | 76.92 | 20.32 | 79.68 |
| DN(70%) + DN | 27.04 | 72.96 | 26.56 | 73.44 | 17.12 | 82.88 | 23.56 | 76.44 |
| DN(70%) + HCW | 22.73 | 77.27 | 27.27 | 72.73 | 15.6 | 84.4 | 19.66 | 80.34 |
| DN(70%) + NF | 29.33 | 70.67 | 28.07 | 71.93 | 20.41 | 79.59 | 23.3 | 76.7 |
| DN(70%) + PF | 30.67 | 69.33 | 40.00 | 60.00 | 28.57 | 71.43 | 21.76 | 78.24 |
| DN(70%) + RC(S) | 24.32 | 75.68 | 30.77 | 69.23 | 18.89 | 81.11 | 21.69 | 78.31 |
| DN(70%) + RC(SE) | 27.86 | 72.14 | 24.44 | 75.56 | 18.45 | 81.55 | 19.35 | 80.65 |
| Pearson chi-square: | 16.7902 | | 9.7281 | | 14.3616 | | 9.3312 | |
| P-Value: | 0.209 | | 0.716 | | 0.349 | | 0.747 | |
