## supplemental table S5 for "When do persuasive messages on vaccine safety steer COVID-19 vaccine acceptance and recommendations? Behavioral insights from a randomised controlled experiment in Malaysia"

**Table S5: Average marginal treatment effects based on interaction with age category with respect to selecting the intent option for definitely accepting the COVID-19 vaccine, and agreeing to recommend the vaccine to healthy adults, elderly, and people with pre-existing health conditions; in each experimental arm relative to control arm.**

| | **Intention to vaccinate** | **Healthy adults** | **Elderly** | **Health condition** |
|---|---|---|---|---|
| | Marginal effects [95% Confidence Interval] | Marginal effects [95% Confidence Interval] | Marginal effects [95% Confidence Interval] | Marginal effects [95% Confidence Interval] |
| **DN(70%)** | | | | |
| Age ≤ 30 | -0.0210 [-0.0685,0.0265] | -0.0357 [-0.0840,0.0127] | 0.0127 [-0.0424,0.0677] | 0.00830 [-0.0570,0.0736] |
| Age > 30 | -0.0289 [-0.0680,0.0103] | -0.0387 [-0.0803,0.00287] | -0.0512* [-0.0974,-0.00510] | 0.0242 [-0.0249,0.0732] |
| **DN** | | | | |
| Age ≤ 30 | 0.0207 [-0.0273,0.0687] | -0.0280 [-0.0750,0.0189] | 0.0131 [-0.0420,0.0681] | 0.0656* [0.00108,0.130] |
| Age > 30 | -0.00321 [-0.0435,0.0371] | -0.0278 [-0.0687,0.0132] | -0.00806 [-0.0532,0.0370] | 0.0873*** [0.0370,0.138] |
| **HCW** | | | | |
| Age ≤ 30 | 0.0121 [-0.0352,0.0593] | -0.0128 [-0.0576,0.0321] | -0.00853 [-0.0631,0.0461] | 0.0249 [-0.0408,0.0906] |
| Age > 30 | -0.0147 [-0.0553,0.0259] | -0.00527 [-0.0462,0.0356] | -0.0349 [-0.0814,0.0116] | 0.0308 [-0.0187,0.0803] |
| **NF** | | | | |
| Age ≤ 30 | -0.00890 [-0.0559,0.0381] | -0.0302 [-0.0778,0.0174] | 0.000910 [-0.0557,0.0575] | 0.0518 [-0.0124,0.116] |
| Age > 30 | -0.0493* [-0.0892,-0.00942] | -0.0218 [-0.0623,0.0186] | -0.0286 [-0.0735,0.0162] | 0.0263 [-0.0228,0.0753] |
| **PF** | | | | |
| Age ≤ 30 | 0.0224 [-0.0256,0.0703] | -0.0513* [-0.102,-0.000985] | 0.00780 [-0.0468,0.0624] | 0.0574 [-0.00624,0.121] |
| Age > 30 | -0.00322 [-0.0428,0.0363] | -0.0104 [-0.0495,0.0286] | -0.0234 [-0.0682,0.0213] | 0.0529* [0.00324,0.103] |
| **RC(S)** | | | | |
| Age ≤ 30 | 0.0192 [-0.0277,0.0661] | -0.0329 [-0.0800,0.0143] | 0.00768 [-0.0447,0.0600] | 0.0432 [-0.0192,0.106] |
| Age > 30 | -0.0186 [-0.0585,0.0212] | -0.0487* [-0.0917,-0.00572] | -0.0504* [-0.0977,-0.00300] | 0.0219 [-0.0282,0.0720] |
| **RC(SE)** | | | | |
| Age ≤ 30 | 0.0184 [-0.0299,0.0668] | -0.0222 [-0.0685,0.0240] | -0.000669 [-0.0546,0.0533] | 0.00916 [-0.0544,0.0728] |
| Age > 30 | -0.0277 [-0.0674,0.0119] | -0.00708 [-0.0467,0.0325] | -0.00515 [-0.0505,0.0402] | 0.0391 [-0.0109,0.0891] |
| **DN(70%)+DN** | | | | |
| Age ≤ 30 | 0.00925 [-0.0376,0.0561] | -0.0237 [-0.0697,0.0222] | 0.00825 [-0.0450,0.0615] | 0.0340 [-0.0293,0.0974] |
| Age > 30 | -0.0249 [-0.0643,0.0145] | -0.0386 [-0.0809,0.00374] | -0.0125 [-0.0565,0.0315] | 0.0432 [-0.00711,0.0935] |
| **DN(70%)+HCW** | | | | |
| Age ≤ 30 | 0.0138 | -0.0163 | 0.0221 | 0.0166 |

| | | | | |
|---|---|---|---|---|
| | [-0.0333,0.0610] | [-0.0612,0.0285] | [-0.0300,0.0741] | [-0.0460,0.0792] |
| Age > 30 | -0.0281 | -0.00652 | 0.00483 | 0.0651* |
| | [-0.0680,0.0117] | [-0.0469,0.0339] | [-0.0392,0.0488] | [0.0149,0.115] |
| **DN(70%)+NF** | | | | |
| Age ≤ 30 | -0.0175 | -0.00655 | -0.0202 | 0.0214 |
| | [-0.0647,0.0298] | [-0.0526,0.0395] | [-0.0768,0.0364] | [-0.0422,0.0850] |
| Age > 30 | -0.0481* | -0.0340 | -0.0388 | 0.0220 |
| | [-0.0883,-0.00788] | [-0.0767,0.00858] | [-0.0850,0.00755] | [-0.0284,0.0724] |
| **DN(70%)+PF** | | | | |
| Age ≤ 30 | -0.00288 | -0.0227 | -0.0520 | -0.0198 |
| | [-0.0507,0.0450] | [-0.0698,0.0244] | [-0.109,0.00516] | [-0.0840,0.0444] |
| Age > 30 | -0.0246 | -0.0261 | -0.0244 | 0.0740** |
| | [-0.0637,0.0145] | [-0.0663,0.0142] | [-0.0695,0.0208] | [0.0252,0.123] |
| **DN(70%)+RC(S)** | | | | |
| Age ≤ 30 | 0.0183 | -0.0155 | 0.0147 | 0.0393 |
| | [-0.0304,0.0671] | [-0.0626,0.0315] | [-0.0412,0.0705] | [-0.0238,0.103] |
| Age > 30 | -0.0152 | -0.0395 | -0.0216 | 0.0463 |
| | [-0.0549,0.0245] | [-0.0813,0.00236] | [-0.0678,0.0246] | [-0.00412,0.0966] |
| **DN(70%)+RC(SE)** | | | | |
| Age ≤ 30 | 0.00311 | -0.0113 | -0.0203 | 0.0187 |
| | [-0.0448,0.0510] | [-0.0580,0.0353] | [-0.0764,0.0358] | [-0.0452,0.0826] |
| Age > 30 | -0.0322 | -0.0235 | -0.0375 | 0.00169 |
| | [-0.0723,0.00791] | [-0.0657,0.0187] | [-0.0845,0.00940] | [-0.0482,0.0516] |
| *N* | 5784 | 5784 | 5784 | 5784 |
