## supplemental table S6 for "When do persuasive messages on vaccine safety steer COVID-19 vaccine acceptance and recommendations? Behavioral insights from a randomised controlled experiment in Malaysia"

**Table S6: Average marginal treatment effects based on interaction with sex with respect to selecting the intent option for definitely accepting the COVID-19 vaccine, and agreeing to recommend the vaccine to healthy adults, elderly, and people with pre-existing health conditions; in each experimental arm relative to control arm.**

| | **Intention to vaccinate** | **Healthy adults** | **Elderly** | **Health condition** |
|---|---|---|---|---|
| | Marginal effects [95% Confidence Interval] | Marginal effects [95% Confidence Interval] | Marginal effects [95% Confidence Interval] | Marginal effects [95% Confidence Interval] |
| **DN(70%)** | | | | |
| Male | -0.0306 | -0.0352 | -0.0149 | 0.0333 |
| | [-0.0743,0.0132] | [-0.0800,0.00960] | [-0.0663,0.0364] | [-0.0228,0.0894] |
| Female | -0.0208 | -0.0419 | -0.0371 | 0.00432 |
| | [-0.0626,0.0211] | [-0.0869,0.00317] | [-0.0866,0.0124] | [-0.0507,0.0593] |
| **DN** | | | | |
| Male | 0.00891 | -0.0228 | 0.00913 | 0.0677* |
| | [-0.0358,0.0536] | [-0.0658,0.0202] | [-0.0423,0.0605] | [0.0120,0.123] |
| Female | 0.00378 | -0.0347 | -0.00717 | 0.0942** |
| | [-0.0390,0.0466] | [-0.0799,0.0106] | [-0.0547,0.0403] | [0.0379,0.151] |
| **HCW** | | | | |
| Male | -0.0313 | -0.0203 | -0.00954 | 0.0320 |
| | [-0.0747,0.0122] | [-0.0649,0.0242] | [-0.0605,0.0415] | [-0.0231,0.0872] |
| Female | 0.0271 | 0.00283 | -0.0362 | 0.0273 |
| | [-0.0176,0.0719] | [-0.0391,0.0447] | [-0.0857,0.0133] | [-0.0295,0.0841] |
| **NF** | | | | |
| Male | -0.0461* | -0.0397 | -0.00518 | 0.0449 |
| | [-0.0900,-0.00210] | [-0.0844,0.00497] | [-0.0564,0.0461] | [-0.0109,0.101] |
| Female | -0.0205 | -0.0114 | -0.0290 | 0.0277 |
| | [-0.0626,0.0216] | [-0.0545,0.0316] | [-0.0774,0.0195] | [-0.0268,0.0823] |
| **PF** | | | | |
| Male | -0.00465 | -0.0202 | 0.0124 | 0.0736** |
| | [-0.0489,0.0396] | [-0.0646,0.0242] | [-0.0386,0.0634] | [0.0179,0.129] |
| Female | 0.0177 | -0.0312 | -0.0320 | 0.0391 |
| | [-0.0244,0.0598] | [-0.0743,0.0119] | [-0.0793,0.0153] | [-0.0160,0.0942] |
| **RC(S)** | | | | |
| Male | -0.0254 | -0.0382 | -0.0137 | 0.0401 |
| | [-0.0688,0.0181] | [-0.0822,0.00578] | [-0.0651,0.0376] | [-0.0152,0.0955] |
| Female | 0.0190 | -0.0476* | -0.0318 | 0.0266 |
| | [-0.0235,0.0614] | [-0.0947,-0.000469] | [-0.0801,0.0164] | [-0.0285,0.0817] |
| **RC(SE)** | | | | |
| Male | -0.00755 | -0.00113 | 0.0411 | 0.0580* |
| | [-0.0521,0.0370] | [-0.0428,0.0406] | [-0.00898,0.0912] | [0.00182,0.114] |
| Female | -0.0114 | -0.0260 | -0.0429 | 0.00147 |
| | [-0.0535,0.0307] | [-0.0700,0.0181] | [-0.0914,0.00563] | [-0.0536,0.0565] |
| **DN(70%)+DN** | | | | |
| Male | -0.0286 | -0.0287 | -0.00707 | 0.0323 |
| | [-0.0717,0.0144] | [-0.0719,0.0146] | [-0.0576,0.0435] | [-0.0231,0.0876] |
| Female | 0.00629 | -0.0375 | -0.00104 | 0.0521 |
| | [-0.0359,0.0485] | [-0.0836,0.00851] | [-0.0466,0.0446] | [-0.00375,0.108] |
| **DN(70%)+HCW** | | | | |
| Male | -0.0337 | -0.0243 | 0.0155 | 0.0233 |

| | | | | |
|---|---|---|---|---|
| | [-0.0771,0.00977] | [-0.0673,0.0187] | [-0.0340,0.0651] | [-0.0324,0.0790] |
| Female | 0.0118 | 0.00619 | 0.00999 | 0.0698* |
| | [-0.0309,0.0545] | [-0.0364,0.0488] | [-0.0357,0.0557] | [0.0149,0.125] |
| **DN(70%)+NF** | | | | |
| Male | -0.0583** | -0.0305 | -0.0407 | 0.0326 |
| | [-0.102,-0.0150] | [-0.0741,0.0131] | [-0.0931,0.0117] | [-0.0230,0.0882] |
| Female | -0.00992 | -0.0138 | -0.0172 | 0.0155 |
| | [-0.0542,0.0344] | [-0.0612,0.0336] | [-0.0669,0.0325] | [-0.0406,0.0716] |
| **DN(70%)+PF** | | | | |
| Male | -0.0272 | -0.0252 | -0.0229 | 0.0452 |
| | [-0.0710,0.0165] | [-0.0686,0.0183] | [-0.0743,0.0285] | [-0.00948,0.1000] |
| Female | -0.00577 | -0.0271 | -0.0465 | 0.0322 |
| | [-0.0475,0.0360] | [-0.0707,0.0165] | [-0.0957,0.00279] | [-0.0231,0.0875] |
| **DN(70%)+RC(S)** | | | | |
| Male | -0.0134 | -0.0229 | 0.00517 | 0.0673* |
| | [-0.0572,0.0304] | [-0.0664,0.0207] | [-0.0458,0.0561] | [0.0125,0.122] |
| Female | 0.00870 | -0.0418 | -0.0178 | 0.0234 |
| | [-0.0346,0.0520] | [-0.0877,0.00418] | [-0.0678,0.0322] | [-0.0329,0.0797] |
| **DN(70%)+RC(SE)** | | | | |
| Male | -0.0412 | -0.00707 | -0.0366 | 0.0129 |
| | [-0.0855,0.00299] | [-0.0501,0.0359] | [-0.0895,0.0162] | [-0.0425,0.0683] |
| Female | 0.00501 | -0.0320 | -0.0217 | 0.00761 |
| | [-0.0381,0.0481] | [-0.0785,0.0145] | [-0.0709,0.0275] | [-0.0482,0.0634] |
| *N* | 5784 | 5784 | 5784 | 5784 |
