## supplemental table S7 for "When do persuasive messages on vaccine safety steer COVID-19 vaccine acceptance and recommendations? Behavioral insights from a randomised controlled experiment in Malaysia"

**Table S7: Average marginal treatment effects based on interaction with education level with respect to selecting the intent option for definitely accepting the COVID-19 vaccine, and agreeing to recommend the vaccine to healthy adults, elderly, and people with pre-existing health conditions; in each experimental arm relative to control arm.**

| | **Intention to vaccinate** | **Healthy adults** | **Elderly** | **Health condition** |
|---|---|---|---|---|
| | Marginal effects [95% Confidence Interval] | Marginal effects [95% Confidence Interval] | Marginal effects [95% Confidence Interval] | Marginal effects [95% Confidence Interval] |
| **DN(70%)** | | | | |
| Below tertiary | -0.0209 | -0.00277 | -0.0429 | 0.000846 |
| | [-0.0633,0.0214] | [-0.0465,0.0410] | [-0.0917,0.00600] | [-0.0547,0.0564] |
| Above tertiary | -0.0305 | -0.0792*** | -0.00658 | 0.0287 |
| | [-0.0739,0.0129] | [-0.126,-0.0322] | [-0.0590,0.0458] | [-0.0270,0.0844] |
| **DN** | | | | |
| Below tertiary | 0.00112 | -0.0113 | 0.00700 | 0.0726* |
| | [-0.0430,0.0453] | [-0.0572,0.0346] | [-0.0417,0.0557] | [0.0165,0.129] |
| Above tertiary | 0.0123 | -0.0455* | -0.00643 | 0.116*** |
| | [-0.0311,0.0558] | [-0.0875,-0.00349] | [-0.0565,0.0436] | [0.0574,0.174] |
| **HCW** | | | | |
| Below tertiary | -0.0143 | 0.0163 | -0.0415 | 0.00298 |
| | [-0.0569,0.0284] | [-0.0269,0.0595] | [-0.0911,0.00803] | [-0.0539,0.0599] |
| Above tertiary | 0.0125 | -0.0336 | -0.00318 | 0.0502 |
| | [-0.0337,0.0587] | [-0.0770,0.00974] | [-0.0543,0.0479] | [-0.00489,0.105] |
| **NF** | | | | |
| Below tertiary | -0.0315 | 0.00302 | -0.0186 | 0.00534 |
| | [-0.0743,0.0114] | [-0.0406,0.0466] | [-0.0673,0.0300] | [-0.0503,0.0610] |
| Above tertiary | -0.0342 | -0.0567* | -0.0179 | 0.0618* |
| | [-0.0776,0.00920] | [-0.101,-0.0121] | [-0.0690,0.0333] | [0.00695,0.117] |
| **PF** | | | | |
| Below tertiary | 0.00972 | -0.0128 | -0.0214 | 0.0183 |
| | [-0.0321,0.0515] | [-0.0566,0.0311] | [-0.0683,0.0255] | [-0.0357,0.0724] |
| Above tertiary | 0.00348 | -0.0358 | 0.00157 | 0.0895** |
| | [-0.0420,0.0490] | [-0.0796,0.00802] | [-0.0507,0.0538] | [0.0314,0.148] |
| **RC(S)** | | | | |
| Below tertiary | 0.00604 | -0.0377 | -0.0459 | -0.0181 |
| | [-0.0363,0.0484] | [-0.0849,0.00951] | [-0.0950,0.00314] | [-0.0737,0.0375] |
| Above tertiary | -0.0138 | -0.0446* | 0.00356 | 0.0815** |
| | [-0.0576,0.0301] | [-0.0881,-0.00114] | [-0.0473,0.0544] | [0.0263,0.137] |
| **RC(SE)** | | | | |
| Below tertiary | -0.00230 | 0.00582 | -0.0144 | -0.00624 |
| | [-0.0453,0.0407] | [-0.0381,0.0498] | [-0.0634,0.0345] | [-0.0626,0.0501] |
| Above tertiary | -0.0177 | -0.0326 | 0.00795 | 0.0585* |
| | [-0.0612,0.0258] | [-0.0742,0.00897] | [-0.0415,0.0574] | [0.00354,0.113] |
| **DN(70%)+DN** | | | | |
| Below tertiary | -0.0179 | -0.0219 | -0.0159 | -0.0126 |
| | [-0.0596,0.0238] | [-0.0667,0.0228] | [-0.0629,0.0312] | [-0.0686,0.0434] |
| Above tertiary | -0.000547 | -0.0382 | 0.00924 | 0.0929** |
| | [-0.0451,0.0440] | [-0.0832,0.00671] | [-0.0403,0.0588] | [0.0373,0.148] |
| **DN(70%)+HCW** | | | | |
| Below tertiary | -0.00594 | 0.0155 | 0.0235 | 0.0310 |

| | | | | |
|---|---|---|---|---|
| | [-0.0502,0.0383] | [-0.0273,0.0583] | [-0.0240,0.0711] | [-0.0257,0.0877] |
| Above tertiary | -0.0151 | -0.0380 | 0.00278 | 0.0597* |
| | [-0.0569,0.0268] | [-0.0810,0.00503] | [-0.0447,0.0503] | [0.00578,0.114] |
| **DN(70%)+NF** | | | | |
| Below tertiary | -0.0414 | -0.0117 | -0.0723** | -0.0245 |
| | [-0.0847,0.00202] | [-0.0582,0.0349] | [-0.124,-0.0209] | [-0.0805,0.0314] |
| Above tertiary | -0.0287 | -0.0353 | 0.0162 | 0.0694* |
| | [-0.0722,0.0149] | [-0.0786,0.00793] | [-0.0346,0.0670] | [0.0129,0.126] |
| **DN(70%)+PF** | | | | |
| Below tertiary | -0.00792 | 0.00264 | -0.0333 | 0.0307 |
| | [-0.0512,0.0354] | [-0.0412,0.0465] | [-0.0840,0.0173] | [-0.0254,0.0868] |
| Above tertiary | -0.0243 | -0.0555* | -0.0372 | 0.0428 |
| | [-0.0666,0.0180] | [-0.0987,-0.0122] | [-0.0874,0.0130] | [-0.0115,0.0970] |
| **DN(70%)+RC(S)** | | | | |
| Below tertiary | 0.00694 | -0.000504 | -0.00328 | 0.00893 |
| | [-0.0370,0.0509] | [-0.0447,0.0437] | [-0.0531,0.0465] | [-0.0479,0.0657] |
| Above tertiary | -0.0113 | -0.0660** | -0.0113 | 0.0776** |
| | [-0.0543,0.0317] | [-0.111,-0.0205] | [-0.0623,0.0397] | [0.0230,0.132] |
| **DN(70%)+RC(SE)** | | | | |
| Below tertiary | -0.0162 | -0.00923 | -0.0552* | -0.0343 |
| | [-0.0596,0.0271] | [-0.0548,0.0364] | [-0.106,-0.00451] | [-0.0904,0.0217] |
| Above tertiary | -0.0193 | -0.0253 | -0.00203 | 0.0497 |
| | [-0.0633,0.0247] | [-0.0691,0.0185] | [-0.0534,0.0494] | [-0.00570,0.105] |
| *N* | 5784 | 5784 | 5784 | 5784 |
